# Drastic changes in collaboration networks and publication patterns in research using the CDC WONDER dataset

**DOI:** 10.64898/2026.01.13.26343992

**Authors:** Danny Maupin, Tulsi Suchak, Abhijit Sengupta, Mariana Marra, Nophar Geifman, Matt Spick

## Abstract

The growth of generative AI and easily available Open Access health datasets has transformed researcher productivity, leading to an explosion in publications that has in part been attributed to paper mills (organisations that provide manuscripts for payment) and other unethical actors. These entities are not, however, homogenous, and have a range of products and target markets. While the demand from China has received much attention, here we provide a case study of CDC WONDER, a dataset that has been exploited by a network of researchers reporting affiliations in Pakistan, the United States and the UK, potentially linked to medical residency driven demand from junior clinicians or trainees. The number of publications using CDC WONDER grew from 88 in 2021 to 1223 in 2025. Over the same time period, the proportion of papers reporting at least one author from Pakistan grew from 0.5% in 2021 to 27.2% in 2025, with unusually extensive collaboration networks. In some cases these works featured over 15 co-authors, often including representation from Western institutions, but in spite of this high level of resourcing only resulted in straightforward analyses of well-described conditions using publicly available data. The majority of these outputs additionally show evidence of being produced from a template, with formulaic titles and identical methods, for example using the same statistical model and platform (Joinpoint regression). Identifying papers produced by fast-churn workflows is essential to protect the integrity of the scientific literature from being flooded with low-quality research. This can be achieved through more proactive desk rejection of misleading and formulaic mass-produced submissions, and through better understanding of which use cases are appropriate for different Open Science resources. With the growing capabilities of AI to mass produce research, education will be essential to assist critical appraisal and preserve the benefits of Open Science.

## Introduction

Generative AI is transforming productivity in data analysis at the same time as Open Science is enabling access to large datasets that any researcher can utilise for hypothesis testing or exploratory research. (1) These innovations democratise science, with benefits ranging from increased access for low- and middle-income countries, the advancement of citizen science, better reproducibility, and outputs being made available for future investigations (2,3). Open Data can, however, be exploited by paper mills, organisations that produce manuscripts for sale. (4,5) This can result in questionable research practices such as data dredging (searching large datasets for any and all possible relationships regardless of whether they are meaningful), HARKing (hypothesizing after the results are known) and p-hacking (selecting variables to maximise statistical significance), often with the goal of mass-manufacturing research outputs.

Misuse of Open Data can also occur when well-intentioned researchers engage in epistemic trespassing, where an expert in one field produces research where they lack expertise and competence (6). Examples of this phenomenon are present throughout the literature, whether they are specific examples of an individual commenting on a separate field (e.g., Linus Pauling a Nobel Prize winner in chemistry arguing for Vitamin C as a treatment for cancer (6)), or within-field trespassing across broader groups such as vaccine hesitancy among health professionals with comments on reliability of vaccine studies and vaccine safety (7,8). This may be done purposefully as part of a preconceived bias, or accidentally due to a lack of knowledge and ability to identify confounding factors.

We have previously described the exploitation of open access datasets through unethical practices; for example the National Health and Nutrition Examination Survey (NHANES) has seen an exponential rise in formulaic and redundant publications (9,10). This trend has also been seen for databases such as the WHO’s Global Burden of Disease Study or the more access-restricted UK Biobank (11), and also with topics that can be applied to multiple datasets, such as Mendelian randomization (12). This exploitation has many harmful effects including: a) overwhelming an already overburdened peer-review system; b) flooding the literature with low-quality outputs, thus undermining the trust in data sources; (13) and c) disseminating low-quality or misleading findings to the wider public.

Despite their significant output, paper mills can be difficult to detect when they use real-world statistics and sophisticated machine learning methods. One possible avenue to aid in their detection is through authorship patterns. Questionable research practices relating to authorship have long been an issue in academia, including adding a large number of authors to bias citations (14) or gift authorship, providing authorship to an individual who did not make a meaningful contribution. (15) However, the metadata from paper mill outputs may provide avenues for their identification. Warning signs might include outliers in author connections (e.g., publications with a large number of institutions) or one-and-done collaborations, where authors collaborate on one paper only with no prior or subsequent outputs, potentially indicating authors that have purchased their place on a paper. Network analysis has previously been employed to investigate such patterns in author relationships and citations for paper mills, for example in finding unusual collaboration patterns at university or country level in the case of a Russia-based paper mill. (16) Author network analysis, however, has not yet been applied to mills that operate in the Open Science data-dredging space.

Given these harmful impacts and adversarial adaptations, it is imperative that new paper mill targets are identified promptly, and that new methods and approaches are implemented to identify unethical actors in the research space. CDC WONDER (or the Centers for Disease Control and Prevention Wide-ranging Online Data for Epidemiologic Research) is a US-provided dataset granting researchers, public health professionals, and the general public access to a wide array of health-related data collections. (17) As with other datasets that have been exploited, it is fully open-access and has been available for interrogation on the Internet since 1995. (18) It is also an example of a dataset that has more recently shown explosive growth in manuscript submissions, possibly indicative of unethical behaviour. In this work we aim to investigate whether the increase in CDC WONDER publications reflects paper mill activity, and whether the growth in submissions carries unusual or changing author network structures.

## Methods

Here, OpenAlex was used as the publication database; OpenAlex was searched with the following string “CDC WONDER”, limited to title/abstract, between the dates of 1 January 1996 and 22 December 2025. OpenAlex was selected for its relational graph capabilities, allowing for automated network analysis, and use of author and institution identifiers. (19)

The resulting dataset of publications was assessed for the prevalence and growth of formulaic and redundant publications using a previously described approach. (9,10) The corpus of publications examined here was split by year, with publications up to 2022 (inclusive) acting as a control group predating the public release of GenAI, and publications from 2023 and onwards acting as a “case” group post the release of GenAI. To help quantify the rise of publications in the CDC WONDER dataset, AutoRegressive Integrated Moving Averages (ARIMA) were used to forecast the expected trend of publications for the control dataset (publications prior to 2023). ARIMA is a time series forecasting model that contains autoregression (uses past values), differencing (accounting for seasonality and other trends), and moving averages (to reduce noise). By comparing the difference between expected production of manuscripts from ARIMA to the actual observed production, an excess production of manuscripts is calculated. The ARIMA model used parameters of autoregressive order *p* = 1, degree of differencing *d* = 1 and moving average order *q* = 1. Confidence intervals were also constructed for the forecasts. ARIMA models were implemented in Python using the statsmodels library. (20) As a second analysis of trend towards growth in formulaic research, token usage in titles was analysed, by simple tokenisation of the published titles and a comparison made of titles in 2025, using the CountVectorizer functionality from the scikit-learn library (21) in Python. Sudden growth in excess of previous trends (that cannot be explained by new interest or data releases) combined with an increase in formulaic or redundant titles has previously been associated with possible paper mill activity.

Network analysis and visualisation was conducted using VOSviewer (22) to identify patterns across countries, institutions and/or authors contributing to the increased use of the CDC WONDER dataset. While country and institution data is presented by name, any author-related information is presented anonymously. A thesaurus file was created to reduce redundant institution names, noting that not all redundancies may have been accurately captured given the range of universities and institutions (Supplemental Table S1).

Lastly, while a full audit of all included papers was beyond the scope of this work, an audit of 40 randomly sampled manuscripts was completed to assess methodological, analytical, or other repetitive similarities.

## Results

An OpenAlex search of works using CDC WONDER was conducted on December 22^nd^, 2025, and returned 2123 publications. Of the 2123, 457 were published prior to 2023 and 1666 articles were published from 2023 and onwards. The vast majority of publications since 2023 were in journals from two publishers: Lippincott Williams (611 articles, 36.7%) and Elsevier (530 articles, 31.8%). No other publishing company accounts for more than 5% of articles published since 2023 (Supplemental Tables S2A and S2B). Figure 1 displays the number of publications and rate of change across journals with 20 or more articles in our dataset.

**Figure 1.**
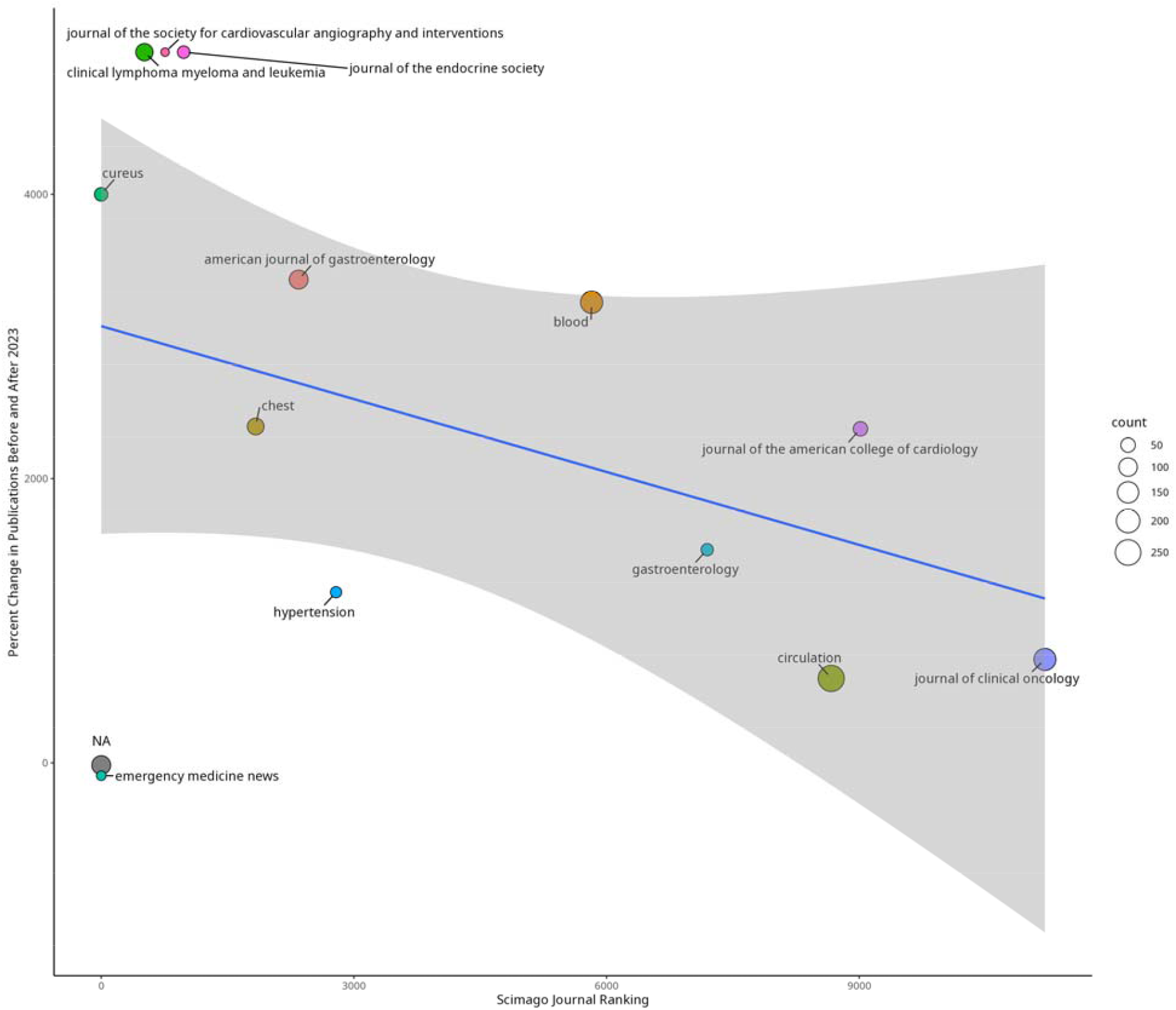
Bubble plot of publication count (size of bubble) and percent change by journal ranking. The journals *Clinical Lymphoma, Myeloma, and Leukemia, Journal of the Endocrine Society, and Journal of the Society for Cardiovascular Angiography and Interventions* have an undefined percent change due to no publications prior to 2023. These have been arbitrarily set to 5000 percent change to highlight their increased use since 2023.

The results of the ARIMA analysis (Figure 2A) indicate a significant deviation from the expected trend as the recorded number of manuscripts falls well outside the 95% confidence interval of the expected number. Patterns in token usage (Figure 2B) showcase a large increase in the terms “trends” and “disparities”, indicative of a formulaic approach to manuscript construction. Given the wide range of years, age / sex groupings, and medical conditions, this template offers very high potential for mass manufacture of publications.

**Figure 2.**
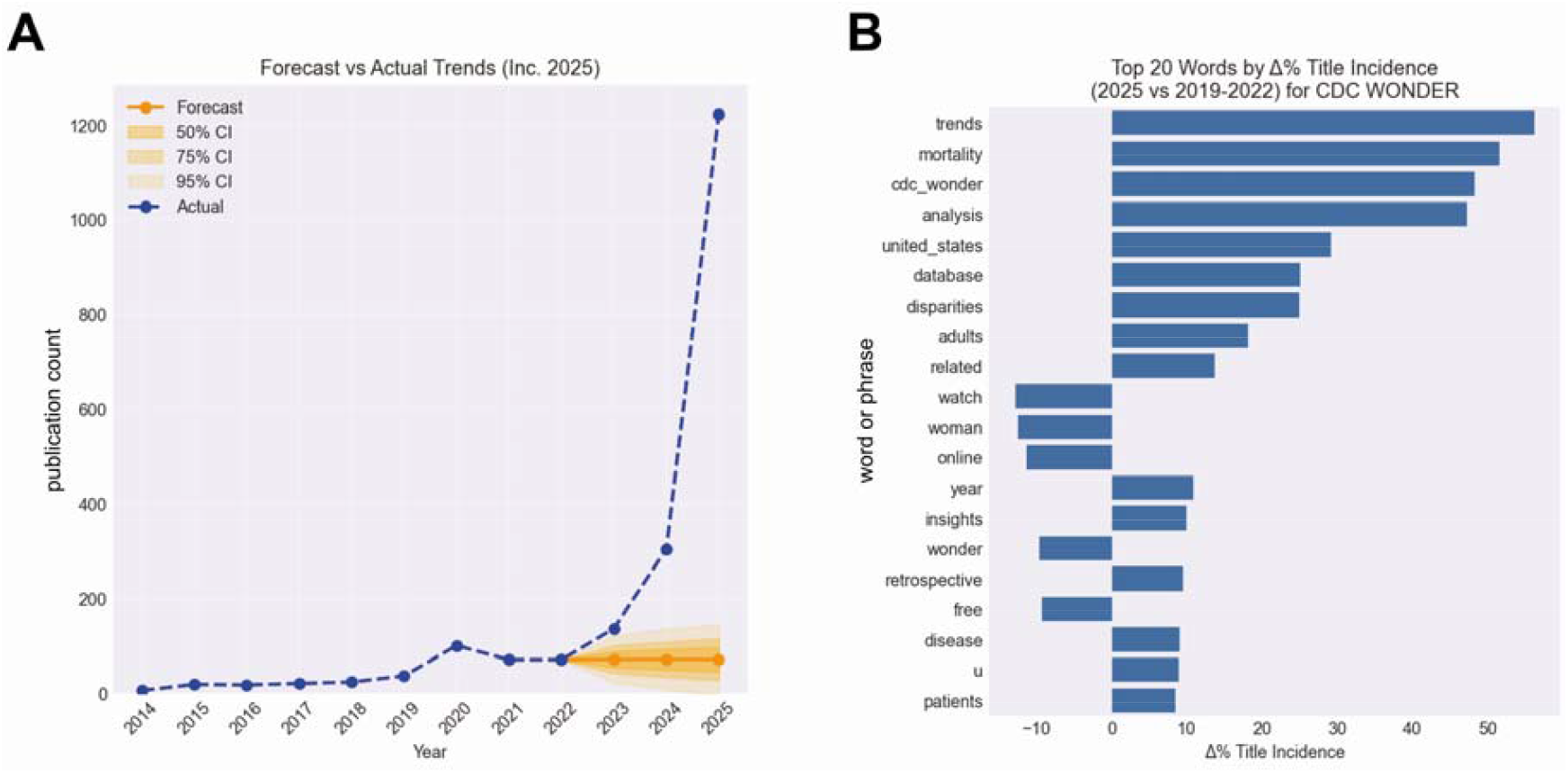
Analysis of trends in publications analysing CDC WONDER data: (A) Annual publication counts and the forecast trend based on the trend up to 2022 (B) changes in tokens in titles of manuscripts

Across the study period, there was a clear upward trend in the average number of authors per paper, increasing from a stable value of two authors per paper, to eight by 2025 (Figure 3). This upward trend is driven, in part, by some papers reporting as many as 31 contributing authors (23).

**Figure 3.**
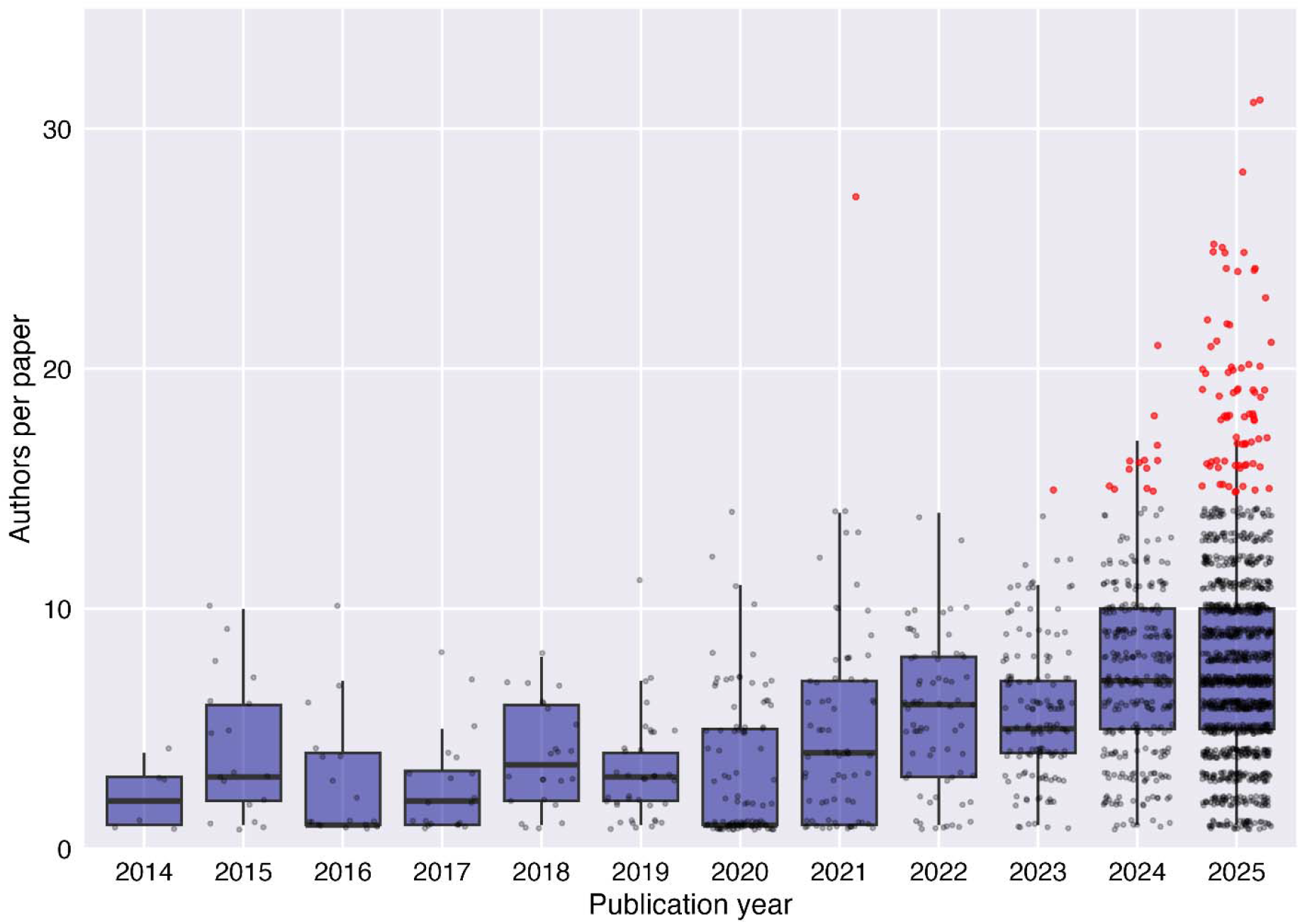
Boxplot of annual average author patterns in publications analysing CDC WONDER data. Red dots indicate papers with more than 20 authors.

Network analysis identified two trends. First, assessing authors’ affiliated country (Figure 4) shows a large number of publications from Pakistan, India, and the U.S. Before 2023, the UK was the highest co-authored country with U.S. authors (with 5.6% of U.S. authors publishing alongside a UK affiliated author). During this same period 0.7% of U.S. authors published alongside an author from either India or Pakistan. Since 2023, 52.7% of U.S. authors have co-authored with an author from Pakistan, 13.2% with an author from India, and 5.7% with an author from the UK, demonstrating significant patterns of contributions with Pakistan and India. In fact, after Egypt with 4.9% of authors, no other country co-occurs with more than 2% of U.S. authors post 2023.

**Figure 4.**
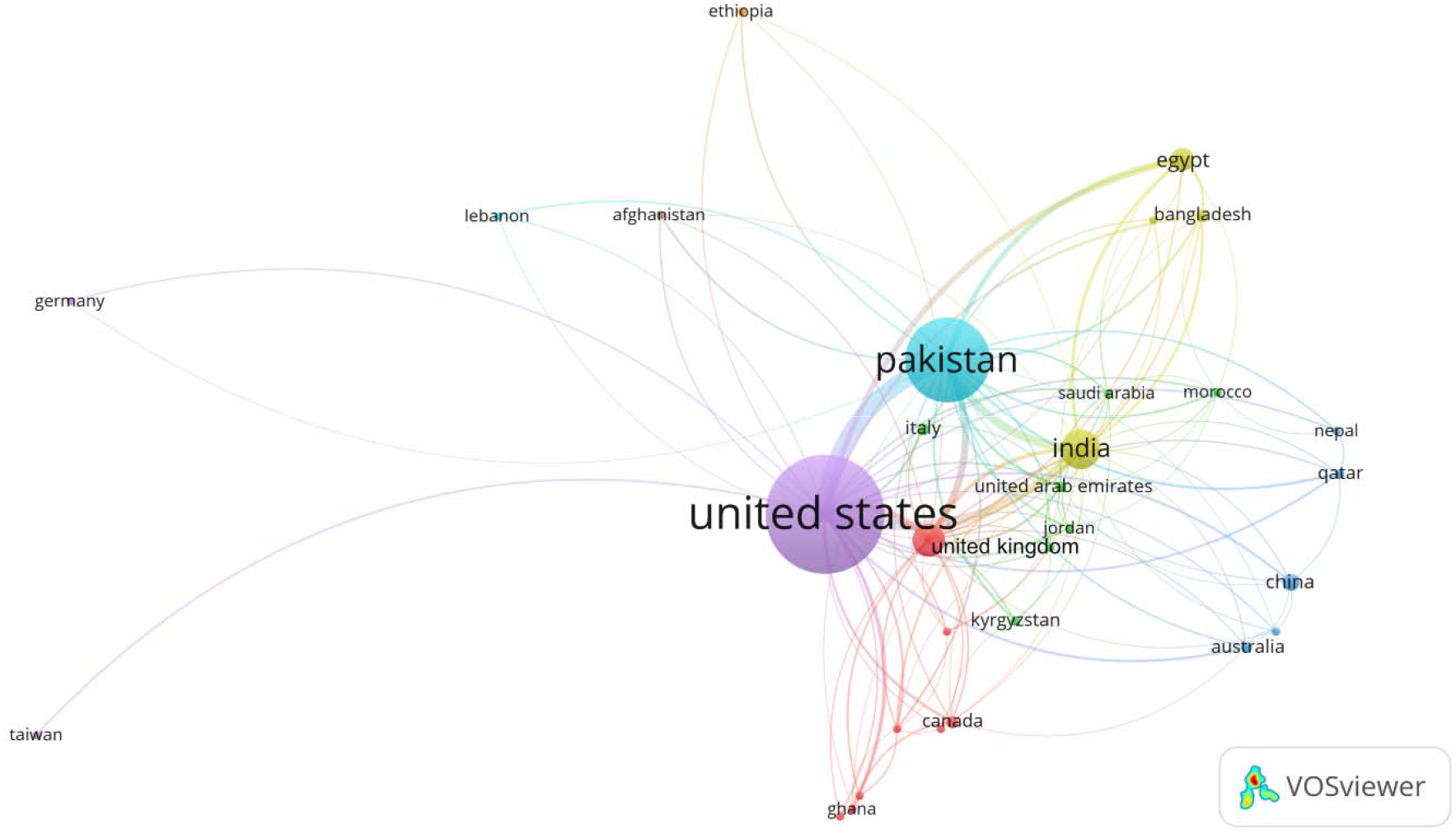
Network analysis of affiliations for publications by country. Colors are applied by a clustering algorithm to aid interpretation using VOSviewer defaults.

**Figure 5.**
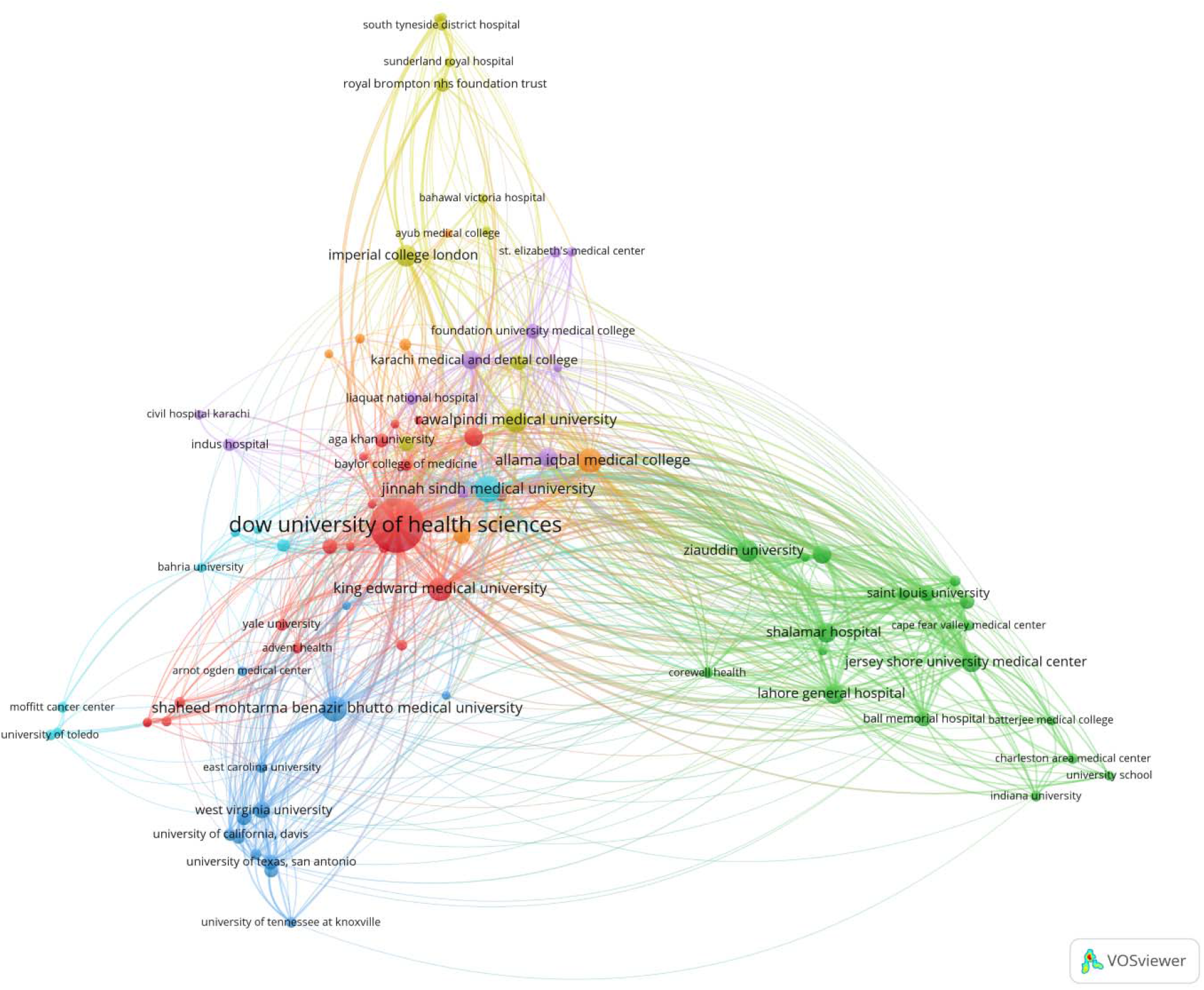
Network analysis of affiliations for publications with at least one author at Dow University of Health Sciences. Colors are applied by a clustering algorithm to aid interpretation using VOSviewer defaults.

The number of countries by author is displayed in Table 1, with a notable increase in authors from Pakistan post 2023, following the pattern of papers with a majority of authors from countries of the Global South with a minority of authors from institutions in the UK and the US.

**Table 1.**
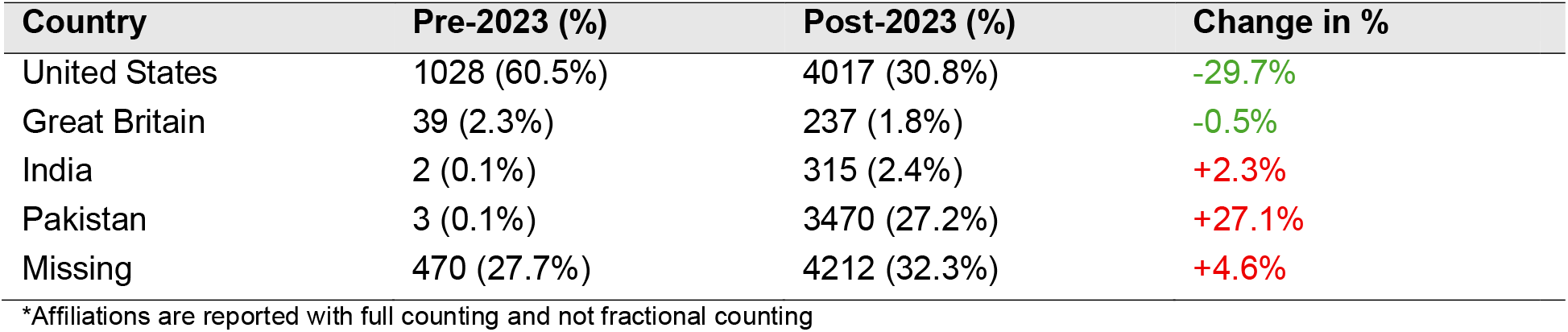
Frequency and proportion of author countries* split into overall, pre-2023, and post-2023.

| Country | Pre-2023 (%) | Post-2023 (%) | Change in % |
| --- | --- | --- | --- |
| United States | 1028 (60.5%) | 4017 (30.8%) | -29.7% |
| Great Britain | 39 (2.3%) | 237 (1.8%) | -0.5% |
| India | 2 (0.1%) | 315 (2.4%) | +2.3% |
| Pakistan | 3 (0.1%) | 3470 (27.2%) | +27.1% |
| Missing | 470 (27.7%) | 4212 (32.3%) | +4.6% |

\*Affiliations are reported with full counting and not fractional counting

Second, an affiliation network for the largest single institutional contributor of CDC WONDER publications (Dow University of Health Sciences, Pakistan) showed a number of clusters, driven by overall linkage strength (number of edges). While some clusters were geographically localised to Pakistan, other clusters included collaborations with the United States (for example one cluster included Duke University as well as Jersey Shore University Medical Centre) and the UK (where a cluster included Imperial College London in addition to hospitals in Sunderland, London and South Tyneside).

Lastly, our random sample of 40 papers found that the same statistical methods and software (Joinpoint regression analysis using NCI software (24)) were used, all had identical data segmentation approaches, and all had essentially identical limitations sections, in spite of having completely different author lists.

## Discussion

This paper provides evidence that CDC WONDER is the latest Open Access data source to be systematically targeted by researchers engaged in questionable or fraudulent research practices. Since 2023, robust evidence has emerged that Open Access data sources are being targeted by unethical researchers and/or paper mills, possibly in combination with GenAI being used to efficiently generate analysis and manuscripts. Articles analysing CDC WONDER have primarily been published with journals owned by Elsevier, Lippincott Williams, and Wiley, including in the form of conference abstracts. This may be indicative of specific targeting of venues with lower barriers by unethical researchers. (25) Further, the inflation of author counts and institutional affiliations per paper (26) could be consistent with paper mill activity and other questionable authorship practices. Whilst the greater number of authors per paper could be seen as a sign of collaboration between institutions, such a stark increase raises questions as to whether all authors met ICMJE authorship requirements. (27) The shift to highly formulaic titles, methodologies and article construction (e.g. identical limitations paragraphs) has also been notable in the last two years. Examples include ‘*Trends and Disparities in Coronary Artery Disease and Obesity-Related Mortality in the United States From 1999–2022*’ and ‘*Trends in Cardiovascular Mortality in Patients With Chronic Kidney Disease From 1999 to 2020: A Retrospective Study in the United States*’. These are examples of manuscripts which follow an identical structure, use the same software (Joinpoint), and provide the same subgroup analyses, which may be indicative of a production-line approach to the research process. (28,29)

In combination, these issues are strongly suggestive of systematic targeting of CDC WONDER to produce fast-churn science. Together with the exponential growth in publication counts, CDC WONDER is the latest Open Science dataset to fall victim to the template-driven, redundant publication approach seen in NHANES and elsewhere (9,11). While the growth in NHANES saw a large increase in publications by authors from China (9), CDC WONDER publications are dominated by authors reporting affiliations from Pakistan and India, typically alongside a small number of authors with an affiliation to a UK or US institution. While this could be authentic international collaboration, the networking patterns of UK or US authors co-authoring publications with very large numbers of collaborators from Pakistan / India, may carry risks of authors gaming the system to add prestige and help pass peer review. Another possible explanation is individuals aiming to connect with Western institutions and authors to add credibility (possibly via training courses led by senior researchers, including those that generate publications using a template at the end of the course). Such activities would naturally be attractive to individuals wanting to use publications to enhance visa applications or acceptance / progress at, for example, medical schools. Recent editorials have highlighted how medical students and residents in the United States, for example, are engaged in research practices that focus on quantity rather than quality. (30,31) This practice adds to the deluge of poor-quality research and potentially undermines public trust in health research and practitioners. Hadidi et al. (31) note that this proliferation in medical school-related research demand can impact conferences in particular; these venues often operate a ‘pay for acceptance’ model with registration fees providing a guaranteed outcome for abstract submissions. Given the network analysis presented here, it is likely that students and residents are targeting Open Access datasets such as CDC WONDER in this way, given the large number of conference submissions to journals such as *Chest* found in our results. We welcome feedback and additional context from any authors that have published a CDC WONDER paper matching these descriptions, especially those that represent genuine international research collaborations.

A number of limitations in this work should be highlighted. The first and most important is that this is an overall analysis of trends, not individual papers, which in some cases may be well-intentioned international collaborations. Second, the results presented here do not represent a full systematic review of the literature. While searching OpenAlex identified approximately 2000 papers, searching Google Scholar, a less restrictive database, using the search terms “trends” AND “disparities” and “CDC WONDER” resulted in 7210 results, over three times that of OpenAlex. This suggests that our analysis may understate the true scale of the problem. Third, a further limitation of this and other works investigating paper mills and other unethical actors is the dependence on keyword searching (here CDC WONDER). Unusual citation or authorship patterns (such as a core of repeating authors, with other co-authors ‘cycled in’ on formulaic outputs) may offer a better way to ‘diagnose’ paper mill activity through network analysis, and represents a promising area of future research across health data outputs. Furthermore, in some cases, publishers have become more restrictive in accepting papers using Open Science data (e.g., PLOS and Frontiers (13)), which creates pressure for paper mills to move on to other sources. Such adversarial adaptations may explain the rapid growth in CDC WONDER exploitation in 2025, but may also lead to a self-correcting reduction in 2026. Nonetheless, this arms race illustrates the tension between Open Science and safeguarding of literature integrity, and reminds us of the importance of active monitoring of these trends in usage. (32)

## Conclusions

We have previously described the tension between Open Science and the need for safeguarding the integrity of the research literature. This is for the simple reason that while open datasets are driven by good intentions, large quantities of freely available data can contribute to AI-driven fast-churn research, and ultimately lead to the credibility of these datasets being questioned. The exploitation of the CDC WONDER dataset provides another case study of this phenomenon. It also demonstrates that such formulaic usage is not limited to specific regions, as the geographic sources of these manuscripts are different to those of NHANES and others, with unusually complex and extensive collaboration networks across Pakistan, the UK and the United States. By identifying the common features of these formulaic and low value manuscripts including unusual collaboration networks and author count inflation, we hope that the publishing community (editors and peer reviewers) will find it easier to reject these types of submissions and avoid both dilution of the scientific literature and also harms to the goals of Open Science.

## Declarations

### Ethics Approval and Consent to Participate

No patients or members of the public were directly involved in this study as no primary data were collected.

### Data availability

The data underpinning this work can be reproduced from the OpenAlex database at https://openalex.org/ from the search strings set out in Methods.

### Author Contributions

MS, AS and NG designed the study, developed the methodology, provided resources, and supervised the project. TS and DM jointly produced the original draft; MS created the visualizations. MS and NG reviewed and edited the manuscript. All authors read and approved the final manuscript. MS is the guarantor of this manuscript and is responsible for the overall content. The corresponding author attests that all listed authors meet authorship criteria and that no others meeting the criteria have been omitted.

